# Decentralizing care for cutaneous leishmaniasis and other skin diseases in Southern Ethiopia: What are the needs?

**DOI:** 10.1101/2024.09.27.24314478

**Authors:** Dagimawie Tadesse, Saskia van Henten, Sifray Batire, Mehret Techane, Tamiru Shibiru Degaga, Behailu Merdekios, Steven Abrams, Asrat Hailu, Jean-Pierre Van geertruyden, Johan van Griensven, Myrthe Pareyn

## Abstract

**Background:** Cutaneous leishmaniasis (CL) and other skin diseases impose a high burden in Ethiopia, yet underreporting is common due to limited access to diagnostics and treatment. Decentralizing care could improve this situation but may necessitate substantial changes in the healthcare system. This study assessed the available resources, and healthcare professionals’ knowledge and skills across Southern Ethiopia’s healthcare facilities to inform decentralization plans.

**Methodology/Principle findings:** A cross-sectional study was conducted from May to July 2023 in Gamo Zone, South Ethiopia, including visits to 11 health centers, 4 primary hospitals, and 1 general hospital. Available resources were evaluated, and clinical and laboratory staff’s knowledge and skills were tested through questionnaires focused on CL and other skin diseases. Most facilities had equipment for diagnosis and localized treatment. Adequate hospitalization space and necessary equipment for systemic CL treatment were found in 3 out of 4 primary hospitals but none of the health centers. Consumable and drug shortages were common across all facilities. BSc laboratory technologists scored significantly higher than diploma technicians (29 *vs.* 15 out of 39, p<0.001). Clinical staff scores varied significantly across education levels (p=0.007), with clinicians scoring the highest (median 33, IQR 31-36), followed by health officers (median 29, IQR 27-32), BSc nurses (median 28, IQR 16-36) and diploma nurses (median 25, IQR 19-29). Notably, no significant differences in median scores were observed between primary hospitals and health centers for both clinical and laboratory staff.

**Conclusions/Significance:** Decentralizing diagnosis and treatment of common skin diseases and localized CL treatment to health centers appears feasible with facility adjustments and continuous staff training. CL cases requiring systemic treatment should be referred to primary hospitals. Strategic efforts to enhance and maintain skills and tackle supply shortages are crucial for successful decentralization.

**Author Summary:** Cutaneous leishmaniasis (CL) and other skin diseases are common in Ethiopia, although many cases remain unreported due to limited access to treatment. This could be improved by providing care at lower healthcare facilities, which may require adjustments. We examined the available resources and (clinical and laboratory) staff skills to diagnose and treat CL and other skin diseases in eleven health centers, 4 primary hospitals, and 1 general hospital in South Ethiopia. All facilities had basic equipment for diagnosis and localized treatment of skin diseases, including CL. However, only primary hospitals had the space and equipment to treat severe CL cases. Consumable and drug shortages were common at all facilities. Staff training needs were evident, with better scores for staff with a higher education. Since skilled staff were distributed across health centers and primary hospitals, overall performance was similar between healthcare levels.

Decentralizing the diagnosis and localized treatment of skin diseases in health centers is feasible with minor facility improvements and continuous training for healthcare workers. Severe CL cases who need hospitalization should be referred to primary hospitals. Strong strategies should be developed to enhance and maintain knowledge and skills and to tackle supply shortages for successful decentralization.

## Background

Cutaneous leishmaniasis (CL) is a neglected tropical disease that causes an important health burden in low- and middle-income countries, including Ethiopia [1–3]. This parasitic infection manifests in severe skin lesions, resulting in scarring and a profound psychosocial impact [1]. CL mainly affects impoverished communities, especially those residing in mountainous regions bordering the Ethiopian Great Rift Valley. Official reports indicate less than 1,000 cases annually [1,4]. However, evidence from several studies in north and south Ethiopia indicates severe underreporting [2,5,6], which suggests incidence estimates may reach 20,000 to 50,000 cases per year [2–4].

Underreporting is influenced by poor health-seeking behavior. This is due to the inaccessibility of diagnostics and treatment, currently only available at 16 treatment centers country-wide, to which patients may have to travel hundreds of kilometers. These centers provide insufficient treatment capacity to manage all CL patients, illustrated by long waiting lists at treatment facilities for the hospitals’ catchment areas [7]. Other barriers to visiting these specialized facilities include a lack of knowledge about diseases and available treatments, as well as economic constraints [8]. Hence, instead of seeking modern treatment for CL, patients visit traditional healers in their community.

CL is not the only prevalent skin disease in rural Ethiopian communities. Other infectious skin morbidities such as scabies, podoconiosis, leprosy, mycetoma, and various fungal and bacterial infections contribute to the overall high burden of skin diseases. The World Health Organization (WHO) roadmap for Neglected Tropical Diseases (NTDs) 2021-2030 [9] and the Ethiopian third national NTDs strategic masterplan 2021-2025 [3] advice for a decentralized healthcare delivery package. They also advocate for an integrated approach for skin NTDs, to optimize the limited available resources. Furthermore, the Ethiopian strategic plan aims to substantially increase the number of facilities that diagnose (currently 25) and treat (currently 14) CL by 2025 to improve access to patients with a target of 170 diagnostic and 30 treatment facilities for CL by 2030 [3].

Improving access to such services through decentralization necessitates important changes to the healthcare system. To what extent healthcare facilities currently have the required equipment, knowledge, and skills to diagnose and treat patients is unknown. This study assessed the available resources in different levels of healthcare facilities and measured the knowledge and skills of corresponding healthcare professionals. Such information is crucial for understanding at which healthcare-level decentralization would be feasible. It also enables the development of a targeted plan for investments and training to achieve successful decentralization.

## Materials and Methods

### Ethical approval

The study was approved by the Arba Minch University Institutional Research Ethics Review Board (194/23), the Institute of Tropical Medicine Institutional Review Board (1680/23), the University Hospital Antwerp Ethics Review Board (5453/23), and the World Health Organization Regional Office For Africa (WHO AFRO) Ethics Review Committee (2023/4.9 [4–1093]). Approval letters from Zonal and District Health offices were obtained to perform the research. A written administrative authorization letter was sought from the head of each healthcare facility before inspecting the infrastructure. Written informed consent for participation in this study was provided by all healthcare professionals before data collection.

### Study setting

The study was conducted in the Gamo Zone, South Region, Ethiopia. Ethiopia follows a three-tier healthcare system with primary, secondary, and tertiary levels of care [10,11]. The primary healthcare level itself consists of three levels. First are primary hospitals, mostly situated in *woreda* (district) towns, in which general practitioners and a few specialists are found. These are equipped with tools to diagnose and treat patients. Second are health centers found in the *woreda* periphery, where nurses, health officers, and sometimes a general physician are found. Basic laboratory and clinical infrastructure are present here. Each health center is linked to five satellite health posts in *kebeles* (villages), the third and lowest level of the primary healthcare system, managed by a health extension worker (HEW). These HEWs get short training and reside in the community where they provide basic health services. The secondary healthcare system includes a general hospital in zonal (regional) cities, which has sophisticated equipment and skilled medical specialists and acts as a referral center for primary hospitals. The tertiary healthcare system includes specialized hospitals and referral centers for general hospitals.

Staff working in these different healthcare levels also have distinct educational backgrounds. Nurses, laboratory personnel, and pharmacy professionals can have two levels: a Bachelor of Science (BSc, undergraduate) requiring four years for nurses and laboratory personnel, and five years for pharmacy programs; and a diploma (certificate) with three years of education at a lower level [12]. Overall, healthcare workers with the highest qualifications, such as physicians and laboratory technologists, were primarily stationed at the hospital levels in our study area (S1 Table). At health centers, the highest level of clinical staff are health officers, with a few exceptions in health centers where physicians work. Healthcare workers with a diploma certificate were evenly available at primary hospitals and health centers.

A total of 64 primary healthcare facilities (one general, five primary hospitals, and 58 health centers) are present in the study area. The study was conducted in 16 purposely selected healthcare facilities based on CL endemicity previously reported [5,12] as well as accessibility, leading to the selection of 1 general hospital, 4 primary hospitals, and 11 health centers (Fig 1).

**Fig 1:**
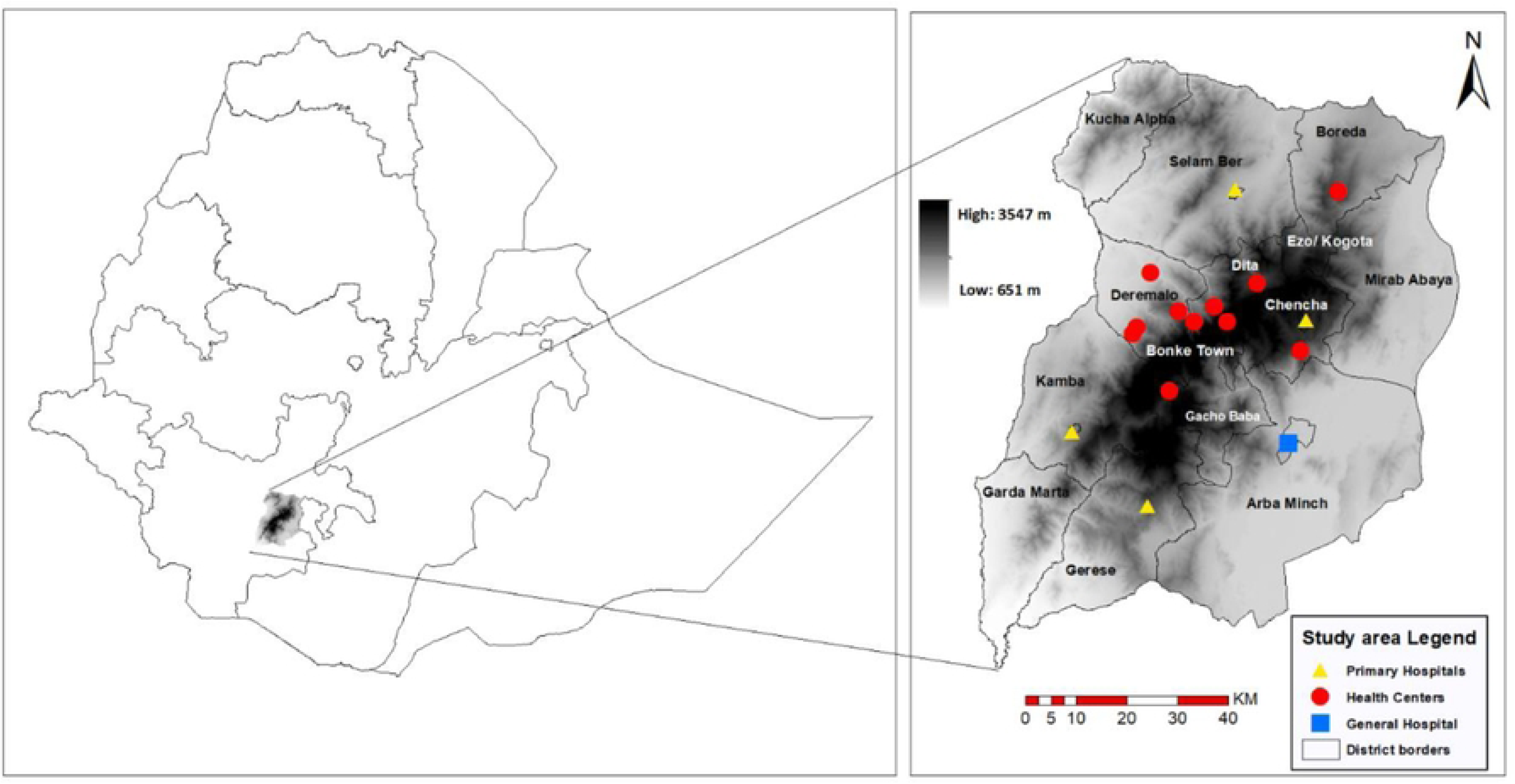
Map showing the healthcare facilities where the study was conducted. **(A)** Map of Ethiopia with study site indicated in the Southern region [13]. **(B)** Zoom map of the study sites on an altitude gradient (14). A blue square displays Arba Minch General Hospital (AMGH); primary hospitals: Chancha, Gerese, Kemba, and Selam Ber are depicted by yellow triangles; and district health centers: Deramalo (Wacha, Guge Boyre, Dara Dime, and Shalladida), Dita (Woyiza, Andiro Giglio, Sulisse, and Zara), Chencha (Dorze), Boreda (Zefine), and Bonke (Gezoso) are indicated by the red circles.

### Study design

A cross-sectional study was performed between May and July 2023 including two components: *(i)* a healthcare facility resource assessment and *(ii)* a laboratory and clinical staff assessment to investigate knowledge and skills for diagnosing and treating CL and other common skin diseases.

### Study population

Comprehensive sampling included all clinical and laboratory professionals at the 16 selected healthcare facilities who *(i)* worked there for over six months regardless of their experience with skin disease diagnosis and treatment, *(ii)* were available during the visit of the study team and *(iii)* provided written informed consent for participation.

### Resource assessment

The selected healthcare facilities were visited by the principal investigator (DT) and a data collector (SB). Information on the catchment population and health insurance was gathered from *woreda* (district) health offices and each facility’s District Health Information System (DHIS). Accompanied by facility staff members and using a predesigned questionnaire, the laboratories, clinical facilities, and pharmacies were visited. The availability of resources such as consumables, permanent infrastructure/tools, and treatments needed to diagnose and manage CL and other common skin diseases were observed. Furthermore, the functionality of equipment (complete blood count (CBC) and chemistry machines, electrocardiograms, beds, etc.) needed for the management of systemic CL treatment was checked; the quality of microscopes was evaluated using Giemsa stained slides from CL patients with a known parasite load by a skilled technologist. The consistency of availability of laboratory consumables (scalpels, microscopy slides, staining reagents, CBC and chemistry reagents, etc.) used for the diagnosis of CL and other skin diseases like Gram stain or potassium hydroxide (KOH) was also recorded.

### Knowledge and skill assessment

At each site, healthcare professionals’ knowledge and skills in diagnosing and treating patients with CL and other skin diseases were assessed, with a primary focus on CL. Novel questionnaires were developed for clinical and laboratory staff separately, based on expert knowledge and discussion within the study team, thereby consisting of a total of 39 multiple-choice questions in each of the questionnaires. The questions were presented on a tablet to each staff member separately in a private room (S2 Table).

More specifically, each questionnaire consisted of 25 knowledge-based and 14 skill-based questions. Clinical staff answered knowledge questions about the signs, symptoms, and treatment for certain diseases. Additionally, they were asked to provide a clinical diagnosis based on lesion pictures to assess their skills (S1 Fig).

Laboratory staff knowledge was assessed through questions about the appropriate diagnostic modalities for different skin diseases. Their skills were evaluated by asking them to identify infectious skin diseases based on microscopic images (S2 Fig). The images employed in both questionnaires were from patients who had previously consented to the anonymized use of their pictures in secondary research, the WHO Skin NTD App, or the Centers for Disease Control and Prevention (CDC) (15,16).

### Data analysis

Data collection was performed using Redcap version 13.8.0 (17), and subsequent cleaning and data analyses were carried out utilizing R Studio version 4.3.2 (18) and ‘ggplot2’ package version 3.5.1 (19) for data visualization. Absolute and relative frequencies (expressed as percentages) were reported for categorical data and medians with corresponding interquartile ranges (IQRs) for continuous variables. When comparing different proportions, corresponding 95% confidence intervals (CIs) were presented. In the knowledge and skill tests, each question received a numerical score for correct (1 point), partially correct (0.5 points), incorrect (0 points), and "does not know" (0 points) responses (S2 Table). The total score based on the questionnaire was computed for each participant by summing up the item-specific scores mentioned previously. Wilcoxon-Mann-Whitney tests were performed to compare the median total scores between two independent groups of participants. When comparing the median score across three or more groups, Kruskal-Wallis tests were used for an overall difference in medians. In the presence of a significant difference between medians (i.e., two-sided p-value <0.05), pairwise comparisons were done using Dunn’s post-hoc test with a Bonferroni correction for multiple testing.

## Results

### Catchment population

The distance of healthcare facilities from Arba Minch General Hospital (AMGH) varies greatly, reaching up to 239 km (Fig 1, and S3 Table). There was a 10-fold difference in the catchment population between the general hospital, primary hospitals, and health centers, and only about half of the population had health insurance (S3 Table).

### Availability of resources

Diagnosis of common skin diseases like CL, scabies, eczema, and fungal and bacterial infections was done in most healthcare facilities. Treatment for these common skin diseases (S4 Table) was available in most facilities, albeit with interruptions. However, rare skin morbidities require referral diagnosis and treatment at higher-level healthcare facilities. CL treatment was available only at the general hospital and sodium stibogluconate (SSG) for intralesional CL treatment was present in one health center, although only available for participants enrolled in an ongoing clinical trial.

For diagnosis of CL, most facilities had a good microscope in place needed to examine Giemsa stained skin slit smears, although supplies of required consumables were unreliable (Table 1).

**Table 1:** Laboratory and clinical capacity availability per healthcare facility level.

| Laboratory Variables |  | Availability | General Hospital<br>N=1 | Primary Hospital<br>N=4 | Health Center<br>N=11 |
| --- | --- | --- | --- | --- | --- |
| Diagnosis of skin diseases | Microscopy space | Available | 1 (100.0%) | 4 (100.0%) | 10 (90.9%) |
|  | Enough staining space | Available | 1 (100.0%) | 4 (100.0%) | 10 (90.9%) |
|  | Microscopy slides | Supply interrupted | - | 4 (100.0%) | 6 (54.5%) |
|  |  | Continuous supply | 1 (100.0%) | - | 5 (45.5%) |
|  | Scalpels | Supply interrupted | - | 1 (25.0%) | 8 (72.7%) |
|  |  | Continuous supply | 1 (100.0%) | - | 1 (9.1%) |
|  | Giemsa | Supply interrupted | - | 2 (50.0%) | 2 (18.2%) |
|  |  | Continuous supply | 1 (100.0%) | 2 (50.0%) | 6 (54.5%) |
|  | KOH | Continuous supply | 1 (100.0%) | 2 (50.0%) | - |
|  | Microscope | Available, but poor quality | - | - | 2 (18.2%) |
|  |  | Good microscope | 1 (100.0%) | 4 (100.0%) | 9 (81.8%) |
| Systemic CL treatment | Hospitalization room | Available | 1 (100.0%) | 4 (100.0%) | 8 (72.7%) |
|  | Functional CBC machine | Available | 1 (100.0%) | 4 (100.0%) | 1 (9.1%) |
|  | Functional Chemistry machine | Available | 1 (100.0%) | 3 (75.0%) | - |
|  | Functional ECG machine | Available | 1 (100.0%) | 3 (75.0%) | - |
|  | CBC reagents | Supply interrupted | - | 2 (50.0%) | - |
|  |  | Continuous supply | 1 (100.0%) | 2 (50.0%) | 1 (9.1%) |
|  | Chemistry reagents | Supply interrupted | - | 3 (75.0%) | - |
|  |  | Continuous supply | 1 (100.0%) | - | - |

Equipment and consumables were split up for diagnosis of skin diseases and systemic treatment of CL. For consumables, three categories were possible: continuous supply, interrupted supply, or no supply (not shown). A dash (-) means no supply (or zero supply) of resources in the healthcare facilities. CBC, complete blood count; ECG, electrocardiogram; KOH, potassium hydroxide. “Reagents needed for the diagnosis of skin disease were shaded light green, while equipment and supplies required for systemic CL treatment were shaded light orange”.

For example, scalpels were available in only one of the 11 health centers (9.1%) and none of the primary hospitals. Giemsa staining was available in about half of the health centers (i.e., 6 out of 11 health centers; 54.5%) and primary hospitals (2 out of 4; 50%).

AMGH and the four primary hospitals had the required spaces for inpatient treatment of patients needing systemic treatment. Although eight out of 11 health centers had a room for patient hospitalization, the bedding capacity and overall quality of the admission spaces were considerably lower at health centers compared to primary hospitals. Chemistry, Complete Blood Count (CBC), and Echocardiographic (ECG) machines - vital for monitoring patients getting systemic treatment for CL - were present in AMGH and three out of four primary hospitals, but were nearly absent in health centers. Chemistry and CBC reagents supplies were also found to be interrupted in most primary hospitals.

### Healthcare professionals experience

A total of 110 healthcare professionals participated in the knowledge and skill assessment, of which 61 (55.5%) were clinical professionals and 49 (44.5%) were laboratory professionals (S1 Table). Despite making up around 60% of the clinical staff in primary hospitals and health centers, only a few diploma nurses were interviewed because of their minor role in direct patient care. In primary hospitals, more physicians (60.0%) were enrolled, while in health centers predominantly health officers (64.5%) were included. Although the trend is similar, proportions are different from the source population.

The median number of years of experience was found to be lower for higher levels of clinical staff, except for specialists (S5 Table). More specifically, physicians showed a median experience of 2 years (IQR 1-3), health officers 3 years (IQR 2-5), and BSc nurses had a median experience of 4 years (IQR 1-12). Laboratory staff had a median of 6 years’ experience (IQR 2-10) for BSc technologists and 5 years (IQR 2-8) for diploma technicians. The median work experience of laboratory professionals was not significantly different between those working in primary hospitals versus health centers (p = 0.378) and between those with a diploma and BSc (p = 0.144; see S3 Fig, panels A and B). Clinical staff working in health centers had significantly more experience (median of 3 years, IQR 2−7) than primary hospital staff (median of 2 years, IQR 1−3, p*=*0.007). No evidence of an overall difference in median experience was observed across different professions at the pre-specified significance level of 5% (p = 0.072; see S3 Fig, panels C and D, respectively). The significant difference in median experience between hospital and health center staff could be slightly driven by a different composition of the staff in terms of profession.

### Questionnaire scores among laboratory professionals

Overall, BSc laboratory technologists outperformed diploma laboratory technicians, with a median total score of 29 (IQR 24−31) and 15 (IQR 9−20) out of 39 respectively (p < 0.001, Fig 2A). Subset analysis showed significant differences in both median knowledge and skills (p < 0.001) sub-scores (S4 Fig, panels A and C) between laboratory professionals’ education levels. The median total scores in primary hospitals (median 20, IQR 14−24) and health centers (median 17, IQR 10−22) did not significantly differ (p = 0.410, Fig 2B). Likewise, there was no notable difference in median knowledge (p = 0.284) and skills (p = 0.769) scores of laboratory professionals between both settings (S4 Fig, panels B, D). Notably, two health centers (Zefine and Wacha) had high-scoring BSc lab technicians with median scores of 29 and 37.

**Fig 2.**
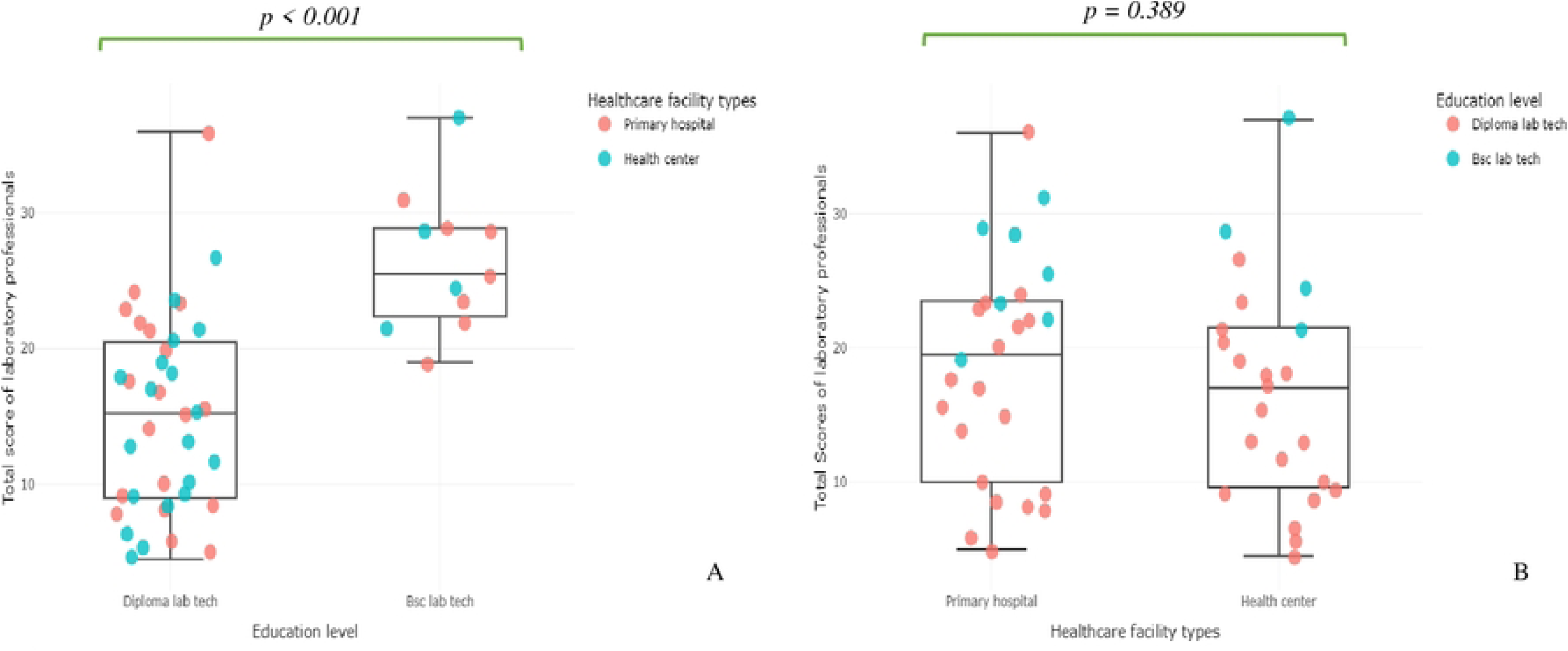
Boxplots showing laboratory professionals’ total knowledge and skill assessment scores. Maximum scores were 39. The scores were compared using Wilcoxon-Mann-Whitney U tests. **(A)** comparisons of the total scores between educational levels of laboratory professionals showed statistically significant differences in median scores (p < 0.001); **(B)** comparison of the median total scores of laboratory professionals in the different healthcare facilities showed no significant difference (p = 0.389).

### Questionnaire scores among clinical professionals

Median total scores on the knowledge and skill assessment combined were significantly different across different educational levels of clinical professionals (p = 0.007, Fig 3A). Diploma nurses had a median score of 25 (IQR 19-29), BSc nurses of 28 (IQR 16−36), health officers of 29 (IQR 27−32), and physicians of 33 (IQR 31−36) (see Fig 3A and S5 Table). Physicians scored significantly better than diploma nurses (adjusted p = 0.026) (Fig 3A) and no differences in median total scores were observed between the other groups. Additionally, in skill questions with lesion pictures (S1 Fig), physicians scored in median terms significantly better than diploma nurses (adjusted p = 0.030) and no significant pairwise differences in medians were observed between other groups. In terms of knowledge, no significant differences in median knowledge scores were observed across groups (S5 Fig, panels A, C).

**Fig 3.**
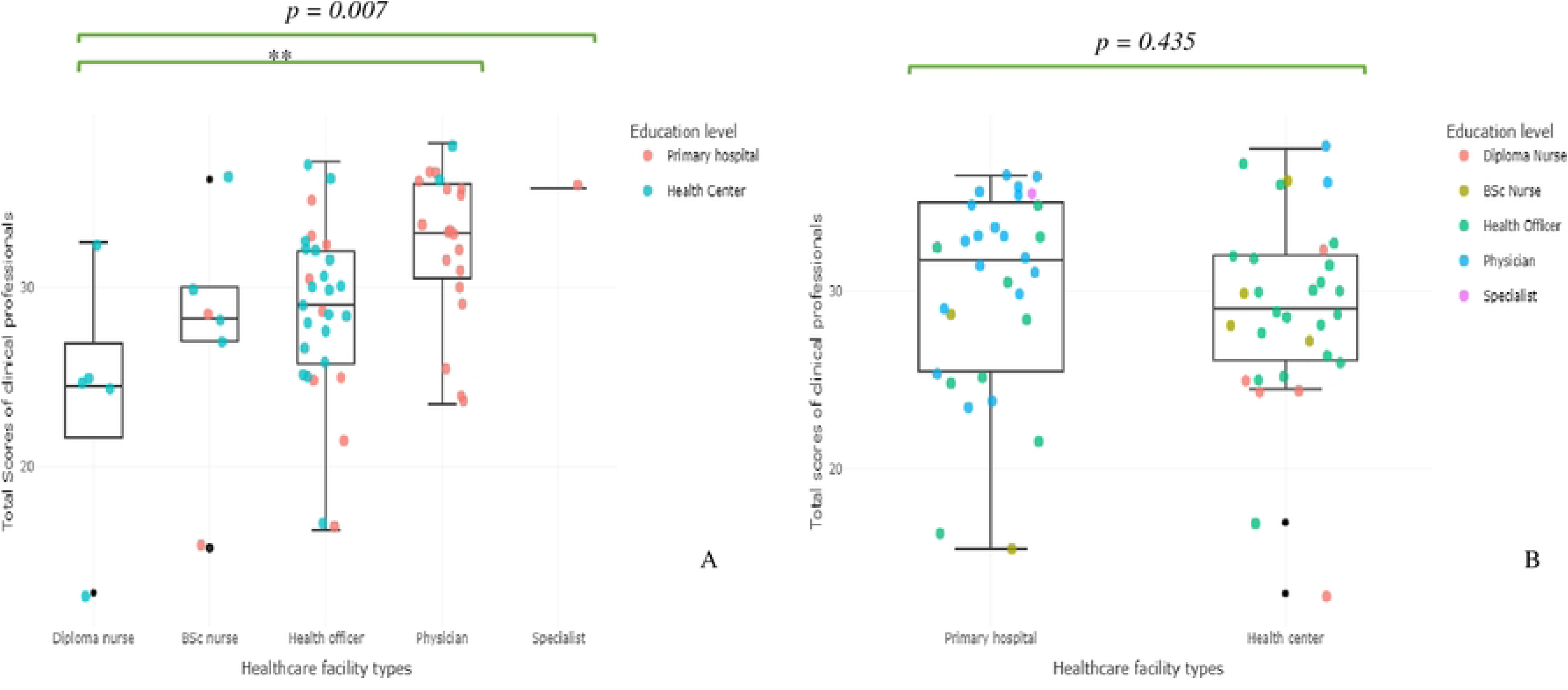
Boxplots showing clinical professionals’ total knowledge and skill assessment scores. Maximum scores were 39 points. The scores were compared using Groupwise (Kruskal-Wallis) and pairwise (Dunn) tests. **(A)** Comparisons of the total questionnaire scores showed a significant difference (p=0.007) in the level of education of clinical professionals (Kruskal-Wallis test). Pairwise (Dunn tests) comparisons with Bonferroni correction showed a significant difference between diploma nurses vs. physicians (adjusted p = 0.026), however, there were no significant differences between health officers vs. physicians (p = 0.079), BSc nurses vs. physicians (p = 0.499), diploma nurses vs BSc nurses (p=1.000), diploma nurses vs health officers (p = 1.000), and BSc nurses vs Health officers (p = 1.000). **(B)** Kruskal-Wallis test comparing the total scores of clinical professionals by healthcare facility showed no significant difference (p = 1.000). ** significance level at *p ≤ 0.05*.

Notably, there was no significant difference (p = 0.435) in the median total scores, as well as for knowledge (p = 0.989) and skill sub-scores (p = 0.076) of clinical professionals between primary hospitals (median 32, IQR 29−33) and health centers (median 29, IQR 27−32) (Fig 3B and S5 Fig, panels B, D). Interestingly, in four health centers (Dorze, Andiro, Zefine, and Wacha) there were clinical staff with scores of 36 or above, amongst which two health officers and two physicians.

## Discussion

Access to care for skin diseases, particularly CL, is poor in Ethiopia. Providing treatment closer to patients could alleviate several significant barriers. Recently, a new treatment center was established in a hospital in Lay Gayint, an area where patients used to have to travel to Gondar more than 200 km away. Following its establishment, over 200 CL patients presented within 20 months (20). According to its strategic plan (3), Ethiopia plans to expand to 30 treatment centers by 2030, making it crucial to understand the necessary human resource and facility investments for sustainability. This study, the first of its kind in East Africa, provides critical data on the current status of healthcare facilities and the knowledge and skills of staff at different healthcare levels in Southern Ethiopia. This information is essential for understanding the feasibility of decentralizing the diagnosis and care for CL and other skin diseases and for identifying the investments required to make this decentralization successful.

### Feasibility of decentralization at different healthcare levels

Our findings highlight a ten-fold increase in the catchment population at each higher healthcare level (S3 Table). A study conducted at Boru Meda Hospital in the Amhara region of Ethiopia demonstrated that long waiting lists of patients can exist at referral centers, leading to delays in receiving systemic CL treatment because their catchment population is so high (21). Given the high estimated number of CL and skin disease patients in the country, decentralization should occur at various lower healthcare levels, supported by an efficient referral system. This approach should prevent higher-level facilities from being overwhelmed by patients.

The overall scores on the knowledge and skills assessments indicate a significant need for further training of both clinical and laboratory healthcare professionals. Although median knowledge and skills scores were considerably higher for those with increasing education levels, no significant differences in median scores were observed between professionals in primary hospitals and health centers. Even though our study population was not equally distributed compared to the source population, this would not have influenced this non-effect. Therefore, our results show that the presence of professionals with higher degrees, rather than the facility level, is crucial for effective decentralization of the diagnosis and treatment of CL and other skin diseases. Of note, both Zefine and Wacha Health centers had high-scoring clinical and laboratory professionals. Therefore, in terms of staff, decentralization can be technically implemented at both primary hospitals and health centers, provided sufficient training is given.

For the diagnosis of CL, rooms for staining and good microscopes were available at almost all primary hospitals and health centers. Thus, clinical diagnosis combined with microscopy for CL could be conducted at both healthcare levels without major resource investments. Once diagnosed, treatment for uncomplicated CL cases can be managed through localized treatments, such as intralesional injections with SSG or cryotherapy. Of note, there were two health centers with knowledgeable lab and clinical staff where skincare could be decentralized, although better availability of skin treatments, as well as supplies for diagnosing CL, need to be guaranteed.

However, many CL patients require systemic treatment, which is currently only available in the form of SSG, which can cause severe renal, cardiac, and hepatotoxicity. Hence, equipment is needed to closely monitor patients before and during treatment. Our study found that chemistry, CBC, and ECG machines, as well as the capacity to hospitalize patients, were present in most primary hospitals but nearly absent in health centers. This indicates that while localized treatment could be provided at both healthcare levels, offering systemic treatment at health centers would require extensive investments in infrastructure and does not seem feasible. Rather, systemic treatment seems more suitable for primary hospitals, where maintenance of the available equipment would still be necessary, and stable supplies (currently not available) need to be guaranteed.

### Addressing challenges

Our study identifies several challenges that need to be addressed to successfully provide care for CL and other skin diseases at lower healthcare levels.

Firstly, frequent stock-outs of reagents for CBC and organ function tests, as well as treatments for various skin diseases and supplies for diagnostic tests such as scalpels, present a significant challenge. Another study conducted in Ethiopia indicated that the supply of SSG is often interrupted (21). Resource shortages were identified as a primary factor affecting the quality of laboratory services in both public and private health facilities in Ethiopia (22). To prevent worsening availability issues if care is extended to additional lower healthcare facilities, it is essential to establish a robust supply management system. Close collaboration between researchers, healthcare providers, and decision- makers will be critical to co-create a strategy for regular staff training and a reliable supply system, making the decentralization of care for skin diseases sustainable.

Another challenge is that about half of the households in Southern Ethiopia have health insurance, which was introduced in 2011 for people living in remote areas (S3 Table). While this is slightly higher than documented previously, where less than a third of the population was insured [24], it still poses challenges in the context of CL. Although SSG is provided free by the Ministry of Health, patients must still pay for laboratory tests and hospitalization, creating a substantial barrier to accessing healthcare. Given that only 50% of patients are cured after one treatment cycle [21,25–27], these costs can accumulate quickly. Increasing insurance coverage to make treatment affordable for all patients and ensuring that hospitals are reimbursed for prefinanced costs is crucial for successful decentralization. This effort will then contribute to achieving Sustainable Development Goal (SDG) 3: equitable access to healthcare (9).

Furthermore, there is a high turnover of healthcare staff, which is reflected by the low number of years of experience, particularly among clinical professionals in primary hospitals. Physicians often leave after a short tenure to pursue residency programs or due to inadequate compensation for duty shifts. A recent meta-analysis confirms a high turnover intention among healthcare providers nationwide (27). Effective retention strategies are crucial to improving the quality of care. In the meantime, frequent retraining of healthcare staff, specifically more stable health officers, will be necessary to ensure accurate diagnosis and quality patient care at lower healthcare levels for CL and other skin diseases. The generalizability of our findings to northern Ethiopia remains uncertain due to differences in infrastructure and healthcare systems. For a larger-scale decentralization intervention across the country, a similar study should ideally be conducted in northern Ethiopia.

## Conclusion

This study indicates that with minor adjustments to facilities and continuous (re)training of primary healthcare professionals, the diagnosis and local treatment of CL may be decentralized to the health center level. Patients with complicated CL requiring systemic treatment should be referred to primary hospitals. Strategic plans with policymakers are essential to address challenges such as low insurance coverage, staff turnover, and stock-outs of reagents and drugs. Proper implementation of an integrated, decentralized care package for skin diseases could significantly enhance access to healthcare for CL and other skin diseases, improve patient outcomes, and reduce the overall disease burden.

## Data Availability

All relevant data are within the manuscript and its files, without restriction.

## Abbreviations

AMGH: Arba Minch General Hospital
BSc: Bachelor of Science
CDC: Centers for Disease Control and Prevention
CBC: Complete Blood Count
CL: Cutaneous Leishmaniasis
DHIS: District Health Information System
ECG: Echocardiographic
HEW: Health Extension Worker
IQR: Interquartile range
KOH: Potassium Hydroxide
NTD: Neglected Tropical Diseases
WHO: World Health Organization

## Acknowledgments

We would like to thank all healthcare professionals who volunteered to participate in this study. We thank the zonal and district health offices and healthcare facilities for their cooperation. We are also grateful to the University of Antwerp biostatisticians Mahdi Safar Pour, Geoffrey Manda, and Tafadzwa Maseko for their support with R studio software training and data analysis.

## Supporting information

S1_fig.docx: Images of skin diseases to assess laboratory professionals’ diagnostic skills.

S2_fig.docx: Images of skin NTD patients used to assess the clinical professionals’ diagnostic skills.

S3_fig.docx: Boxplots showing clinical laboratory professionals’ work experience

S4_fig.docx: Boxplots showing laboratory professionals’ knowledge and skill assessment scores.

S5_fig.docx: Boxplots showing clinical professionals’ knowledge and skill assessment scores.

S1 Table: Descriptive summary of the source and sampled population

S2 Table: Questionnaires used for knowledge and skill assessment

S3 Table: Distance to Arba Minch, catchment population, and health insurance data of study sites

S4 Table: Availability of diagnosis and regular treatment for CL and other common skin diseases

S5 Table: Comparative analysis of clinical and laboratory professionals’ median (and IQR) scores and work experience against different levels of education and healthcare facilities.

## References

1. World Health Organization [Internet]. 2018 [cited 2018 Aug 29]. Leishmaniasis. Available from: http://www.who.int/leishmaniasis/disease/en/.

2. Seid A, Gadisa E, Tsegaw T, Abera A, Teshome A, Mulugeta A, et al. Risk map for cutaneous leishmaniasis in Ethiopia based on environmental factors as revealed by geographical information systems and statistics. Geospatial Health. 2014 May;8(2):377–87.

3. Ethiopian Federal Ministry of Health. The Third National Neglected Tropical Diseases Strategic Plan 2021-2025 (2013/14 – 2017/18 E.C.). November 2021. Available from: https://espen.afro.who.int/system/files/content/resources/Third%20NTD%20national%20Strategic%20Plan%202021-2025.pdf

4. Bugssa G. The Current Status of Cutaneous Leishmaniasis and the Pattern of Lesions in Ochollo Primary School Students, Ochollo, Southwestern Ethiopia. Sci J Clin Med. 2014;3(6):111.

5. Merdekios B, Pareyn M, Tadesse D, Getu S, Admassu B, Girma N, et al. Detection of Cutaneous Leishmaniasis Foci in South Ethiopia. Am J Trop Med Hyg. 2021 May 10;105(1):156–8.

6. Alvar J, Vélez ID, Bern C, Herrero M, Desjeux P, Cano J, et al. Leishmaniasis worldwide and global estimates of its incidence. PloS One. 2012;7(5):e35671.

7. Zewdu FT, Tessema AM, Zerga AA, Henten S van, Lambert SM. Effectiveness of intralesional sodium stibogluconate for the treatment of localized cutaneous leishmaniasis at Boru Meda general hospital, Amhara, Ethiopia: Pragmatic trial. PLoS Negl Trop Dis. 2022 Sep 9;16(9):e0010578.

8. Tamiru HF, Mashalla YJ, Mohammed R, Tshweneagae GT. Cutaneous leishmaniasis a neglected tropical disease: community knowledge, attitude and practices in an endemic area, Northwest Ethiopia. BMC Infect Dis. 2019 Oct 16;19(1):855.

9. WHO. A road map for neglected tropical diseases 2021-2030. 2020. https://www.who.int/teams/control-of-neglected-tropical-diseases/ending-ntds-together-towards-2030/targets.

10. The Ethiopian health care system [Internet]. [cited 2024 Mar 3]. Available from: https://bio-protocol.org/exchange/minidetail?type=30&id=8778625

11. Zebre G, Gizaw AT, Tareke KG, Lemu YK. Implementation, experience, and challenges of urban health extension program in Addis Ababa: a case study from Ethiopia. BMC Public Health. 2021 Jan 19;21:167.

12. Pareyn M, Kochora A, Van Rooy L, Eligo N, Vanden Broecke B, Girma N, et al. Feeding behavior and activity of Phlebotomus pedifer and potential reservoir hosts of Leishmania aethiopica in southwestern Ethiopia. PLoS Negl Trop Dis. 2020;14(3).

13. https://open.africa/dataset/2018-ethiopia-shapefiles [Internet]. [cited 2024 May 23]. Available from: https://africaopendata.org/

14. ArcMap Resources for ArcGIS Desktop | Documentation, Tutorials & More [Internet]. [cited 2024 May 23]. Available from: https://www.esri.com/en-us/arcgis/products/arcgis-desktop/resources

15. WHO - Skin NTDs App | InfoNTD [Internet]. [cited 2024 Mar 5]. Available from: https://www.infontd.org/practical-material/who-skin-ntds-app

16. Search Results | CDC [Internet]. 2022 [cited 2024 Mar 5]. Available from: https://www.cdc.gov/search/index.html

17. Harris PA, Taylor R, Thielke R, Payne J, Gonzalez N, Conde JG. Research electronic data capture (REDCap)--a metadata-driven methodology and workflow process for providing translational research informatics support. J Biomed Inform. 2009 Apr;42(2):377–81.

18. Download R-4.3.2 for Windows. The R-project for statistical computing. [Internet]. [cited 2023 Dec 12]. Available from: https://cran.r-project.org/bin/windows/base/

19. Villanueva RAM, Chen ZJ. ggplot2: Elegant Graphics for Data Analysis (2nd ed.). Meas Interdiscip Res Perspect. 2019 Jul 3;17(3):160–7.

20. Yizengaw E, Nibret E, Yismaw G, Gashaw B, Tamiru D, Munshea A, et al. Cutaneous leishmaniasis in a newly established treatment centre in the Lay Gayint district, Northwest Ethiopia. Skin Health Dis. 2023 Aug;3(4):e229.

21. van Henten S, Bialfew F, Hassen S, Tilahun F, van Griensven J, Abdela SG. Treatment of Cutaneous Leishmaniasis with Sodium Stibogluconate and Allopurinol in a Routine Setting in Ethiopia: Clinical and Patient-Reported Outcomes and Operational Challenges. Trop Med Infect Dis. 2023 Aug 14;8(8):414.

22. Mesfin EA, Taye B, Belay G, Ashenafi A, Girma V. Factors Affecting Quality of Laboratory Services in Public and Private Health Facilities in Addis Ababa, Ethiopia. EJIFCC. 2017 Oct;28(3):205–23.

23. Merga BT, Balis B, Bekele H, Fekadu G. Health insurance coverage in Ethiopia: financial protection in the Era of sustainable cevelopment goals (SDGs). Health Econ Rev. 2022 Aug 3;12(1):43.

24. Fikre H, Mohammed R, Atinafu S, van Griensven J, Diro E. Clinical features and treatment response of cutaneous leishmaniasis in North-West Ethiopia. Trop Med Int Health. 2017 Oct 1;22(10):1293–301.

25. Tesfa D, Manaye N, De Vries HJ, Van Griensven J, Enbiale W. Clinical pattern and treatment outcome of Cutaneous Leishmaniasis in two hospitals in Bahir Dar, Ethiopia (2017-2021). J Infect Dev Ctries. 2022 Aug 31;16(8.1):26S-34S.

26. van Henten S, Tesfaye AB, Abdela SG, Tilahun F, Fikre H, Buyze J, et al. Miltefosine for the treatment of cutaneous leishmaniasis—A pilot study from Ethiopia. PLoS Negl Trop Dis. 2021 May 28;15(5):e0009460.

27. Gebrekidan AY, Enaro EY, Azeze G, Adella GA, Kassie GA, Haile KE, et al. Turnover intention among healthcare workers in Ethiopia: a systematic review and meta-analysis. BMJ Open. 2023 May 1;13(5):e067266.

